# MULTIMODAL-MULTITASK-SELFSUPERVISED XDEEP-MSI: EXPLAINABLE BIAS-REJECTING MICROSATELLITE INSTABILITY DEEP LEARNING SYSTEM IN COLORECTAL CANCER

**DOI:** 10.1101/2022.12.29.22284034

**Authors:** Aurelia Bustos, Artemio Payá, Andres Torrubia, Cristina Alenda

## Abstract

The prediction of microsatellite instability (MSI) in colorectal cancer (CRC) using deep learning (DL) techniques directly from hematoxylin and eosin stained slides (H&E) has been shown feasible by independent works. Nonetheless, when available, relevant information from clinical, oncological and family history could be used to further inform DL predictions. The present work analyzes the effects from leveraging multimodal inputs and multitask supervision in a previously published DL system for the prediction of MSI in CRC (xDEEP-MSI). xDEEP-MSI was a multiple bias rejecting DL system based on adversarial networks trained and validated in 1788 patients from a total of 25 participating centers from EPICOLON and HGUA projects. In the present work, xDEEP-MSI is further enriched with weakly supervised learning in multiple molecular alterations (MSI status, K-RAS and BRAF mutations and Lynch Syndrome confirmed by germline mutations), adapted to multimodal inputs with variable degree of completeness (image, age, gender, localization of CRC, revised Bethesda criteria, Amsterdam II criteria and additional oncological history) and a self-supervised multiple instance learning that integrates multiple image-tiles, to obtain patient-level predictions. The AUC, including all three selected tissues (tumor epithelium, mucin and lymphocytic regions) and 5 magnifications, increases from 0.9 ± 0.03, to 0.94 ± 0.02. The sensibility and specificity reaches 92.5% 95%CI(79.6-98.4%) and 93.4% 95%CI(90.0-95.8%) respectively. To the best of our knowledge this is the first work that jointly uses multimodal inputs, multiple instance learning and multiple molecular supervision for the prediction of MSI in CRC from H&E, demonstrating their gains in performance. Prospective validation in an external independent dataset is still required.

## 1 Introduction

Approximately 3% of colorectal cancers (CRC) arise in the context of Lynch syndrome (LS), where the patient has a germline mutation in a DNA mismatch repair (MMR) gene [1].Historically, CRC patients were tested for LS if they were at high risk according to clinical criteria, e.g. aged under 50 years or with a strong family history. Several clinicopathologic criteria such as Amsterdam criteria, revised Bethesda guidelines, (see Table 4) were used to identify individuals at risk for Lynch syndrome or eligible for tumor-based MSI testing[2]. However, a large proportion of LS patients were missed by this strategy [3]. Currently, new diagnostics guidance are recommending that all patients with newly diagnosed CRC be screened for LS. Universal tumor-based genetic screening for Lynch syndrome, with MSI or IHC testing of all CRCs regardless of age, has greater sensitivity for identification of Lynch syndrome as compared with other strategies [1]. The pathway includes testing tumour tissue for defective MMR(dMMR) by either microsatellite instability (MSI) testing or immunohistochemistry (IHC) for the MMR proteins MLH1, PMS2, MSH2 and MSH6. Tumours showing MSI or MLH1 loss should subsequently undergo BRAF mutation testing followed by MLH1 promoter methylation analysis in the absence of a BRAF mutation. Patients with tumours showing MSH2, MSH6 or isolated PMS2 loss, or MLH1 loss/MSI with no evidence of BRAF mutation/MLH1 promoter hypermethylation, are referred for germline testing if clinically appropriate.

MSI is not specific for LS, and approximately 15 percent of all sporadic CRCs and 5 to 10 percent of metastatic CRCs demonstrate MSI due to hypermethylation of MLH1 [4, 5, 3]. Sporadic MSI-High (MSI-H) CRCs typically develop through a methylation pathway called CpG island methylator phenotype (CIMP), which is characterized by aberrant patterns of DNA methylation and frequently by mutations in the BRAF gene. These cancers develop somatic promoter methylation of MLH1, leading to loss of MLH1 function and resultant MSI. The prevalence of loss of MLH1 expression in CRC increases markedly with aging and this trend is particularly evident in women [6].

While guidelines set forth by multiple professional societies recommend universal testing for dMMR/MSI [7], these methods require additional resources and are not available at all medical facilities, so many CRC patients are not currently tested [8].

Since the last two decades, certain histology-based prediction models that rely on hand-crafted clinico-pathologic feature extraction - such as age <50, female sex, right sided location, size >= 60 mm, BRAF mutation, tumor infiltrating lymphocytes (TILs), a peritumoral lymphocytic reaction, mucinous morphology and increased stromal plasma cells - have reported encouraging performance but has not been sufficient to supersede universal testing for MSI/dMMR [9]. Measurement of histology variables for MSI prediction, requires significant effort and expertise by pathologists, and inter-rater differences may affect the perceived reliability of histology-based scoring systems [9]. However, this work is fundamental to the premise that MSI can be predicted from histology, which was recently proposed as a task for deep learning from digital pathology [10, 11]. Such an automated screening tool could be used in the clinic to triage patients for confirmatory testing, potentially reducing the number of tested patients, thereby resulting in substantial test-related labour and cost savings.

Research on deep learning methods to predict MSI directly from hematoxylin and eosin (H&E) stained slides of CRC have proliferated in the last years, citing among others [10, 11, 12, 13, 14, 15, 16, 17, 18, 19, 20]. A systematic review of the most relevant works is available in [21]. Published systems can be classified in two types, those trained with weak supervision, where only patient-level labels are used to provide supervision signal for labeling large regions of tissue in whole slide and/or TMA spot images, hence introducing noisy labels. [10, 11, 12, 13, 14, 15, 16, 17, 18, 19] vs system trained on image-level/region or pixel-level annotations as in [20] where they performed the registration of MMR IHC stained slides to the target slide.

Among current limitations, in addition to the challenge of noisy labels introduced by weak supervision, the developed systems so far are not able to distinguish between somatic and germline etiology of MSI, such that confirmatory testing is required. Additionally, there is relevant clinical information for the predicition of MSI status that even if commonly available such as age and CRC localization, those are not yet being exploited by DL-based MSI prediction systems. Both clinical and family history information when available for a given patient, could be used to help improve model predictions for MSI status. Also, the majority of DL systems reported were trained only on the final task of predicting MSI status, nonetheless multitask supervision has demonstrated in multiple domains to outperform narrow specialized system trained on single targets. In particular, for CRC, a system trained to learn not only the MSI status but a more wide tumor molecular phenotype including the different pathways and mutations involved, both somatic as well as germline mutations associated with Lynch Syndrome, has not yet been attempted. Of note, even if different works have attempted the prediction of *KRAS mut* and *BRAF V600 mut* directly from H&E stained tumor slides, in particular for *KRAS* performance metrics are still far below 0.8.[22, 19, 23]. Another important limitation is generalizability due to batch effects, while those systems have proved excellent performance on well curated cohorts that are similar to training data, the performance is not robust to differing patient and tissue characteristics.

In this study, we aimed to test if the performance of deep learning (DL)-based models for the prediction of MSI status in CRC can be improved by leveraging multimodal inputs as well as by training them on multitask supervision in multiple molecular alterations, including both somatic and germline mutations. For this end, we consider as baseline the xDEEP-MSI system [18], which already incorporated a multiple bias-ablation DL technique to avoid learning dataset batch-effects, and built upon it, leveraging multimodal inputs consisting of digitalized tumor histopathology images, general clinical variables, oncology and family history variables -with varying degree of completeness- and adding weak multitask supervision in multiple biomarkers. Also, self-supervised multiple instance learning with per patient attention-mechanisms is integrated to prioritize the tiles of tissue regions with high signal-to-noise and overcome the limitations of weak supervision. To systematically demonstrate the gains in performance from each of those approaches the system is trained and validated in consecutive experiments using the same study samples as in xDEEP-MSI. We demonstrate, how each of those new approaches contributes incrementally to improve the results.

The remainder of the paper is as follows. Section 2 describes the methodology employed, including the study population, partitioning methods, deep learning architecture detailing each of the new modules, the training regime, the management of missing and imbalanced data and explainability methods. Section 3 describes the results of the different MSI-prediction models trained in consecutive experiments with increasing input modalities entered and supervision in different tasks. It also describes the results for the prediction of *KRAS mut, BRAF V600 mut* and Lynch Syndrome. Lastly it analyzes the effect of incomplete data at inference time for MSI status prediction. Section 4 addresses the discussion, conclusions and future work.

## 2 Material and Methods

### 2.1 Study Population, Data Collection and Ground Truth Ascertainment

The study population, data collection and ground truth ascertainment is described in detail in [18]. Briefly it consists of 57 tissue-microarrays (TMAs) that included cylindrical H&E stained tissue samples of 1 mm diameter each (spots), in duplicate from all patients prospectively collected in the EPICOLON project. The EPICOLON project was a population-based, observational, cohort study which included 1705 patients with CRC from 2 Spanish nationwide multi-center studies: EPICOLON I [24] and EPICOLON II [25]with the aim of investigating different aspects related to the diagnosis of hereditary CRC. It was assumed that the EPICOLON population was representative of the Spanish population, due to the large number of participating centres (25) being most of them referral centers, their homogeneous distribution throughout the country, and the lack of ethnic differences among regions [24]. Both studies were approved by the institutional review boards of the participating hospitals. The overall MSI frequency in the EPICOLON project was 7.4%.

The population was further expanded with 283 additional patients retrospectively obtained from the *Hospital Universitario de Alicante, Spain* (HGUA) from 2017 to 2020, which -once preprocessed-added 66 MSI-H and 177 microsatellite stable (MSS) cases (34% MSI frequency) to the final study population as shown in Figure 1. The corresponding H&E images were provided in 15 TMAs that included 1 mm spots in duplicate for each patient.

**Figure 1:**
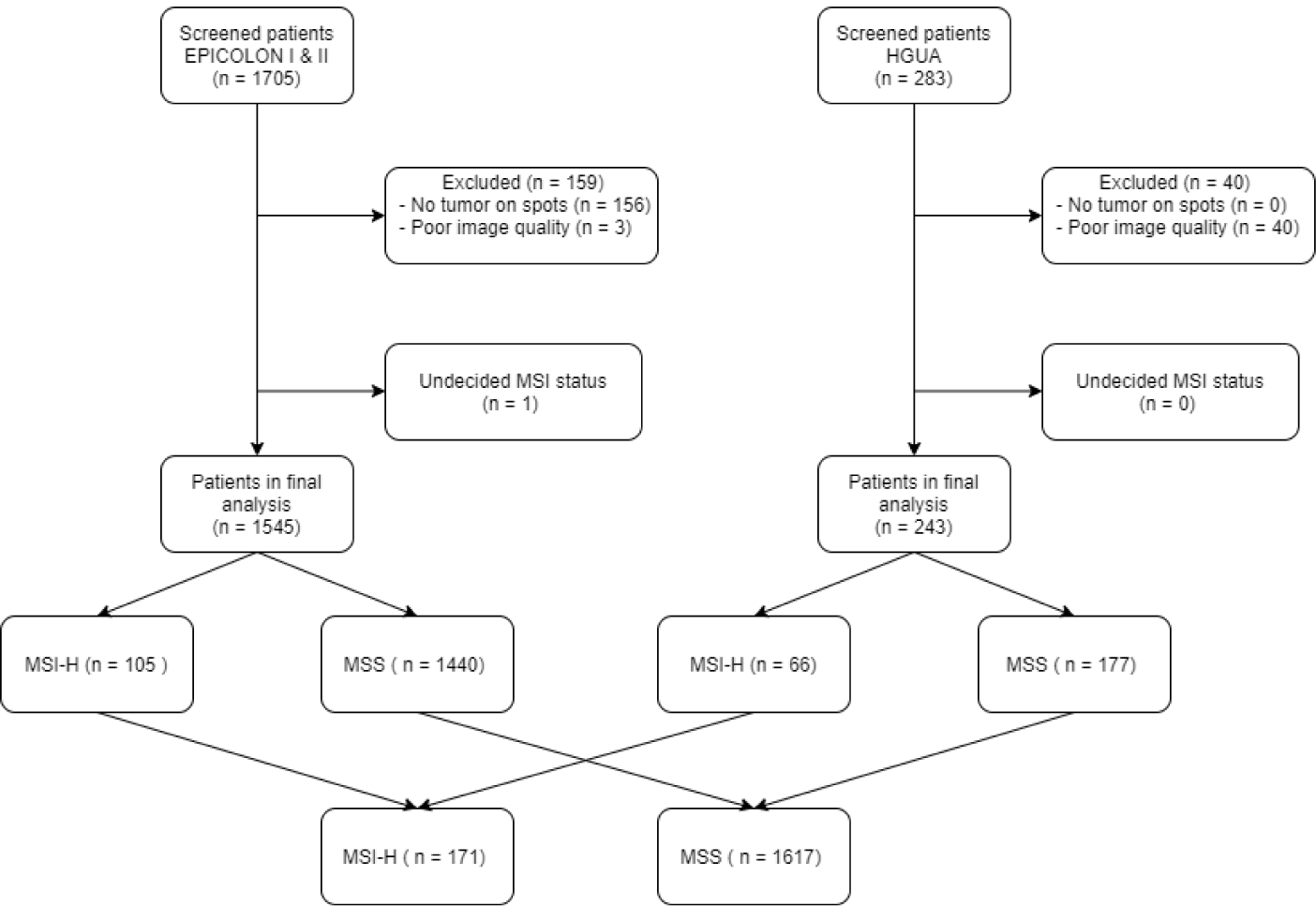
Study sample flowchart: For image preprocessing, the tissue classifier module was used to exclude images based on the criteria of no remaining tumor epithelium on spots.

Patient’s demographic and general clinical characteristics of the final study sample after preprocessing are summarized in Table 1.

**Table 1:** Patient General Characteristics (n = 1788): Main demographic and clinical characteristics of patients included in the final study sample after image preprocessing. The percentage of non-missing characteristics reflects the completeness of variables for the whole study sample. All variables but Duke’s stage (which was available for less than 50% of patients), were used as part of the multi-modal inputs in the system

| Variable | n | Total non-missing (%) |
| --- | --- | --- |
| All patients | 1788 | 1788 (100%) |
| Age (years) | 75.6 (27-101) | 1481 (82%) |
| Gender |  | 1786 (93%) |
| Male | 1037 (58.1%) |  |
| Female | 749 (41.9%) |  |
| Localization primary tumor |  | 1362 (76.2%) |
| Right colon | 393 (28.9%) |  |
| Left colon | 969 (71.1%) |  |
| Dukes’ Stage |  | 891 (49.8%) |
| I | 119 (6.6%) |  |
| II | 370 (20.6%) |  |
| III | 271 (15.1%) |  |
| IV | 131 (7.3%) |  |

The present work additionally includes the tumor molecular phenotype, together with main patient’s oncology and family history variables as available (with varying degree of completeness) from both projects (EPICOLON and HGUA). The tumor molecular characteristics collected in the present work (see Table 2), in addition to the MSI status, included Lynch syndrome confirmed by germline mutation (Yes vs No), *KRAS* mutation (*mut*) vs wild-type (*wt*), *BRAF V600 mut* vs *wt* and a composite variable *KRAS-BRAF* (*KRAS wt* and *BRAF wt* vs *KRAS mut* or *BRAF mut*). This composite variable was introduced to help mimic therapeutic decisions on the metastatic setting where patients are commonly considered for targeted therapy with anti-EGFR factors only if both *KRAS-BRAF* are wild-type.

**Table 2:** Tumor Molecular Alterations (n = 1788): Molecular alterations of CRC tumors analyzed in the final study sample. All variables were used for multitask supervised training as explained for each experiment. Missing values were masked for loss computation. The percentage of non-missing alterations reflects the completeness of molecular variables for the study sample. Wild-type *wt*, Mutation *mut*, Germline mutation *germ-mut*

| Variable | n | Total non-missing (%) |
| --- | --- | --- |
| All patients | 1788 | 1788 (100%) |
| MSI status |  | 1788 (100%) |
| MSI-H | 171 (9%) |  |
| MSS | 1617 (91%) |  |
| *KRAS* |  | 379 (21.2%) |
| *KRAS mut* | 115 (30.4%) |  |
| *KRAS wt* | 264 (69.6%) |  |
| *BRAF* |  | 428 (24%) |
| *BRAF mut* | 40 (9.3%) |  |
| *BRAF wt* | 388 (90.7%) |  |
| *KRAS-BRAF* |  | 370 (20.7%) |
| *KRAS and BRAF wt* | 215 (58.1%) |  |
| *KRAS or BRAF mut* | 155 (41.9%) |  |
| Lynch Syndrome *germ-mut* |  | 1574 (88.0%) |
| Yes | 13 (0.8%) |  |
| No | 1561 (99.2%) |  |

Patient’s oncology and family history variables consisted of main tumor and family history information of patients, relevant for the prediction of both hereditary CRC (Lynch Syndrome) and MSI status, which were collected as part of the EPICOLON project. None of those variables were available in the dataset from the HGUA project. The variables included the 5 Revised Bethesda Criteria described in Table 4, Amsterdam II Criteria [26] (Yes vs No), prior diagnosis of CRC, synchronic diagnosis of CRC, prior diagnosis of adenomatous polyps of colon, synchronic diagnosis of adenomatous polyps of colon, number of family members diagnosed with a CRC, number of family members diagnosed with any related hereditary nonpolyposis colorectal cancer (HNPCC). HNPCC-related tumors include colorectal, endometrial, stomach, ovarian, pancreas, ureter and renal pelvis, biliary tract, and brain (usually glioblastoma as seen in Turcot syndrome) tumors, sebaceous gland adenomas and keratocanthomas in Muir-Torre syndrome, and carcinoma of the small bowel.

The *ground-truth* consisted of labeled spot images as positive or negative for specific genetic mutations or molecular phenotypes without detailed annotations at the cellular and regional levels, which is commonly referred as weakly supervised learning [19]. As described in [18], for the task of MSI prediction each patient and corresponding spots were labeled as MSI-H vs. MSS. MSI-H was defined as tumour-tissue testing defective MMR(dMMR) by either microsatellite instability (MSI) testing or immunohistochemistry (IHC) for the MMR proteins MLH1, PMS2, MSH2 or MSH6. Tumours showing MSI or MLH1 loss underwent *BRAF* mutation testing followed by *MLH1* promoter methylation analysis in the absence of a *BRAF* mutation. Briefly, patients with tumours showing MSH2, MSH6 or isolated PMS2 loss, or MLH1 loss/MSI (regardless of presence or not of *MLH1* promoter hypermethylation), were labeled as MSI-H and all others as MSS. Lynch syndrome germline mutations, *KRAS* and *BRAF V600* testing and categorization methods are described in [24] and [25].

TMAs were scanned with the VENTANA ROCHE iSCAN scanner at magnification ×40 corresponding to a maximal resolution of 0.25 microns per pixel (MPP).

This project was approved by the institutional research committee CEIm PI2019-029 from ISABIAL and both the images and associated clinical information was previously anonymized. The data from EPICOLON project and HGUA are not publicly available, in accordance with the research group and institutional requirements governing human subject privacy protection.

### 2.2 Multimodal System Architecture

The system architecture is a pipeline of integrated technical elements, denoted as modules (see Figure 3), for the prediction of MSI status from CRC H&E-stained tissue image and other multimodal clinical inputs.

**Figure 2.**
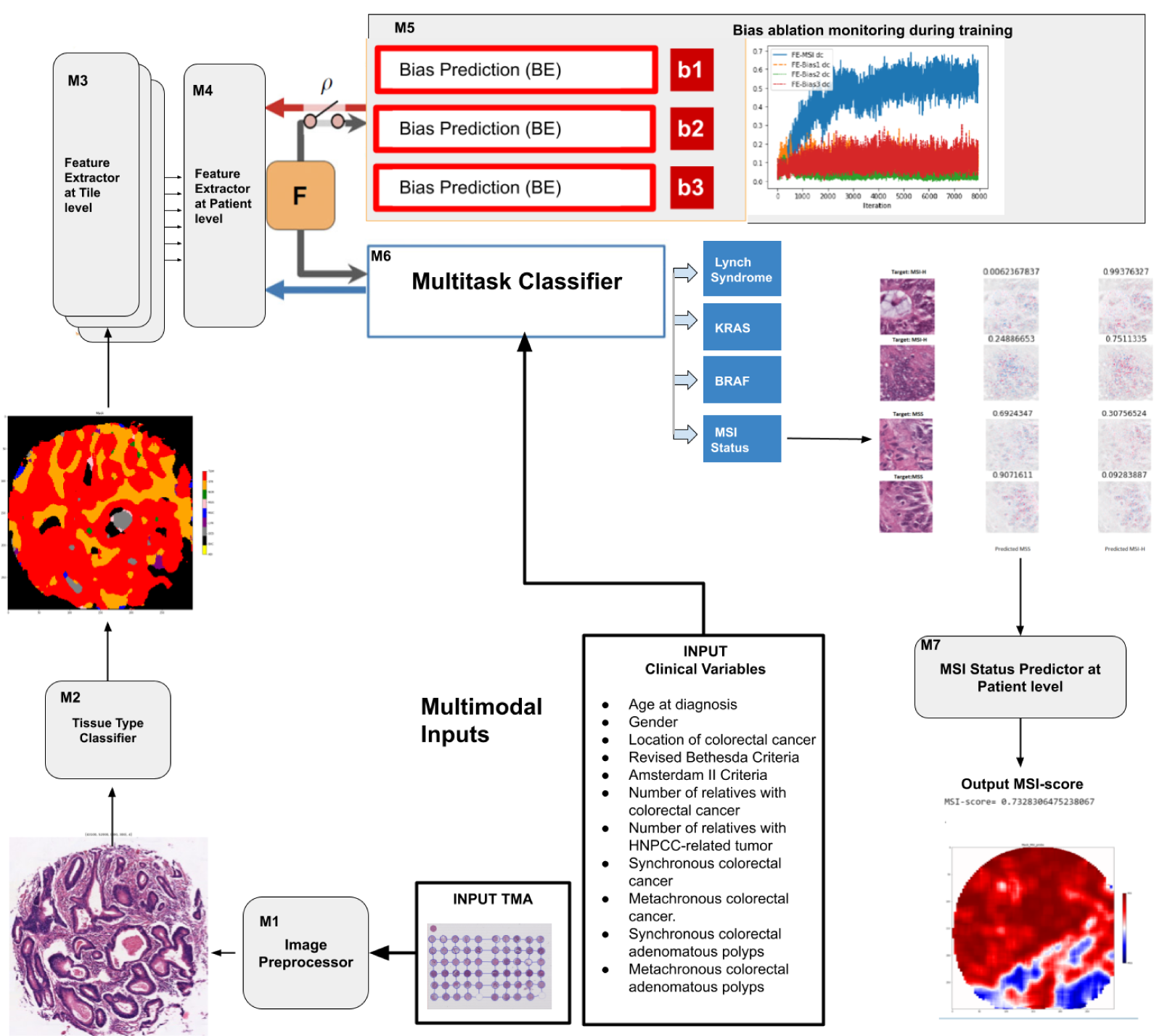

**Figure 3:** System architecture: (**a**) The system is composed of seven modules (M). After spot tilig and pre-processing by M1, M2 classify each tile in 9 tissue types and only tiles corresponding to tumor epithelium, mucine and lymphocitic infiltrates are selected for further processing. M3 extract image features at tile level -which are sampled stochastically for each patient- and then fed into M4, a multiple instance learner that integrate them and extract features at patient level, *F*. M6 encode and concatenate clinical inputs with the patient-level image features *F* to classify the input in the primary outcome *MSI* while being supervised by multiple secondary tasks (Lynch Syndrome, *BRAF* and *KRAS mut*). *F* also remains invariant (statistically independent and conditioned by *ρ*) to the biases responsible for multiple batch effects, *b*_*n*_, using the adversarial components BE and the adversarial loss of M5. The multitask classifier (M6) aims to find the relation to enable prediction of the output labels while the adversarial components (M5) aim to remove the direct dependency between *F* and *b*_*n*_. Figure adapted from [18, 27] by renaming and adding modules and adding multiple adversarial components to the architecture. http://creativecommons.org/licenses/by/4.0/.

#### 2.2.1 M1: Preprocessing Module

The image preprocessing module consists of a TMA-customized dynamic extractor of tiles of programmable size, extracting the corresponding coordinates, mean and standard deviation, corresponding to adjacent regions, without overlap, parameterized at different magnifications (*×*40, *×*20, *×*10, *×*5, *×*0).

#### 2.2.2 M2: Tissue-type Classifier Module

.The tissue classifier module is a pre-trained DL-model (described in [18] which reached an AUC of 0.98 in the validation set) that classifies image tiles obtained by M1 in 9 types of tissues: adipose (ADI), background without tissue (BACK), debris (DEB), lymphocytes (LYM), mucus or mucine (MUC), smooth muscle (MUS), normal colon epithelium (NORM), cancer-associated stroma (STR) and colorectal adenocarcinoma epithelium (TUM). Only spots that contains any region of viable tumor tissue (TUM) are selected. Tiles are subsequently filtered by regions of interest which are restricted to tumor epithelium, lymphocytic infiltrates and mucine, discarding other tiles.

#### 2.2.3 M3: Tile-level Feature Extractor Module

This module reuses the model parameters of M2 as a feature extractor backbone (removing the tissue-type head layers). It receives as input the tiles filtered by the tissue types of interest obtained from M2. The parameters of M3 are retrained end-to-end with subsequent modules. M3 outputs a tile-level image representation of 1024 intermediate features, which serves as input to M4.

#### 2.2.4 M4: Patient-level Feature Extractor Module

This module is a self-supervised multiple instance learning layer based on a encoder-decoder transformer architecture. The dimension of features in the encoder and decoder inputs is 1024, the number of heads in the multihead-attention model is 8 and the number of layers in both the encoder and decoder is 2. The input to this module is the latent spaces, obtained by M3, from a set of *n* tiles for each patient, using a stochastic sampling method that aims to be representative of the different tissue regions found in all spots available for each patient. The attention-mechanisms in M4 in turn are designed to focus on the image tiles with the highest signal-to-noise ratio for each patient with respect to the MSI state. Also, the magnification level of each tile, embedded in 1024 dimensions, is optionally inputted as a query to the decoder. This module finally outputs a patient-level image representation *FE* of 1024 intermediate features, which serve as input to both the M5 and M6 modules.

#### 2.2.5 M5: Multiple Bias Ablation Module

This module applies the method described in [18] to address multiple biases at the DL architecture. Namely it was designed to avoid learning the multiple batch-effects *b*_*n*_ introduced in the image dataset by the study project of origin, the patient and the TMA glass on which the spots were found. Those three factors were shown to be biases that statistically correlated with the MSI-status hence providing an unfair shortcut for MSI status prediction when not properly addressed. The ablation of each bias *b*_*n*_, was implemented through an adversarial training and distillation bias regime following the approach described in [27], and expanded to all three biases identified, following the same architecture as described in [18]. Briefly, each of the bias ablation modules *BE* have two hidden layers of dimension 1024 with ReLU as the activation function, and 2 out features. To monitor the distillation of biases during training, the squared distance correlation *dc* [28] was computed on each batch-iteration between the image features *FE* and the three biases (study project, patient, and glass) and *FE* and the MSI status. The *dc* is a measure of dependence between random vectors analogous to product-moment covariance and correlation, but unlike the classical definition of correlation, distance correlation is zero only if the random vectors are statistically independent [28].

The learned features from the bias-ablated model were verified, in the different experiments done in the present study, to have maximum discriminative power with respect to the task of MSI prediction and minimal statistical mean dependence with the biases.

#### 2.2.6 M6: Multitask Classifier of multiple molecular characteristics

This module has three hidden fully connected layers with 1041, 1024 and 512 dimensions respectively, ReLU as the activation function, and 10 out features to train the model in 5 binary supervised predictive tasks: MSI-status, which is considered the primary task, *BRAF V600 mut, KRAS mut, KRAS or BRAF mut* and Lynch Syndrome. The module receives as inputs the vector representations of all multimodal data (obtained from both image and clinical variables). Specifically the patient-level image representation *FE* of 1024 intermediate features (obtained from M4) is concatenated with the encoded clinical variables that include 1) age at diagnosis, 2) gender, 3) tumor location, 4) revised Bethesda criteria (see Table 4), 5) Amsterdam v2 criteria, 6) number of relatives with colorectal cancer, 7) number of relatives with HNPCC-related tumor**, 8) synchronous colorectal cancer, 9) metachronous colorectal cancer, 10) synchronous colorectal adenomatous polyps, 11) metachronous colorectal adenomatous polyps. Missing input data is dynamically masked during training and inference, allowing different combinations of variables to be inputed in the system depending on the completeness and availability of information for each patient.

#### 2.2.7 M7: MSI Status Prediction Module

This module invokes *m* times module M6 in inference mode applying test-time image augmentations. At each time *m*, a total of *n* pre-filtered tiles are randomly sampled from a given patient hence obtaining a statistical distribution of *mxn* predictions per each patient.

M3,M4,M5 and M6 are trained end-to-end applying at each batch iteration two types of training regimes: 1) a multi-task supervision regime on multiple tumor molecular characteristics, computing a total loss per batch iteration equal to the sum of the task-specific losses, which is back-propagated from M6 to M4 and from M4 to M3 and 2) an adversarial training on *b*_*n*_ which is back propagated from M5 to M4 and from M4 to M3 to distillate *b*_*n*_ from both M4 and M3 as described in [18].

### 2.3 Benchmarks

*xDEEP-MSI*, described in [18] which was trained on image only, was used as a baseline image model to compare results for the models trained in the experiments with multimodal inputs. Additionally a second baseline model, trained on clinical data without image, was implemented to compare results with the multimodal DL-system when only clinical data without image was inputted on inference. This second baseline model was based on gradient boosted decision trees which are currently considered the best performing methods for tabular data and a limited number of observations available. It was implemented using the CatBoost library [29] in the Python programming language. During training, a set of decision trees is built consecutively. Each successive tree is built with reduced loss compared to the previous trees. The number of trees is controlled by the starting parameters. 10-fold cross-validation was applied with the goal to select model hyper parameters. Training parameters shared across all CatBoost models trained were learning rate = 0.3, maximum depth = 6, loss function = Cross Entropy, evaluation metric = AUC. The final model was the best model applying early stopping.

### 2.4 Missing and Imbalanced Data

As shown in Tables 1, 2 and 3, missing data was very frequent depending on the variable. To address this shortcoming, not only at training but also at inference time (which is a common circumstance in the routine clinical practice), both the training regime and predictive models were designed to be resilient to incomplete data.

**Table 3:** Patient’s Oncology and Family History (n = 1788): Main tumor and family history information of patients included in the final study sample collected from the EPICOLON project, relevant for the prediction of both hereditary CRC and MSI status. The percentage of non-missing characteristics reflects the completeness of variables for the whole study sample. All variables were used as part of the multi-modal inputs in the system. Revised Bethesda Criteria are described in Table 4. Amsterdam II Criteria are described in [26]

| Variable | n | Total non-missing (%) |
| --- | --- | --- |
| All patients | 1788 | 1788 (100%) |
| Prior CRC |  | 1165 (65.2%) |
| Yes | 11 (0.9%) |  |
| No | 1154 (99.1%) |  |
| Synchronous CRC |  | 1165 (65.2%) |
| Yes | 64 (5.4%) |  |
| No | 1101 (94.6%) |  |
| Prior colon adenoma |  | 1169 (65.4%) |
| Yes | 24 (20.6%) |  |
| No | 1135 (79.4%) |  |
| Synchronous colon adenoma |  | 803 (44.9%) |
| Yes | 216 (26.8%) |  |
| No | 587 (73.2%) |  |
| Family with CRC |  | 1788 (100%) |
| Yes | 210 (11.7%) |  |
| No | 1578 (88.3%) |  |
| N Family with CCR | 0.56 (0-8) | 462 (25.8%) |
| N Family with HNPCC-related tumor | 1.15 (0-6) | 1165 (65.2%) |
| Revised Bethesda Criteria (RBC) |  | 1165 (65.2%) |
| Yes | 261 (22.4%) |  |
| No | 904 (77.6%) |  |
| RBC 1 |  | 1164 (65.1%) |
| Yes | 42 (3.6%) |  |
| No | 1122 (96.4%) |  |
| RBC 2 |  | 1165 (65.2%) |
| Yes | 88 (7.5%) |  |
| No | 1077 (92.5%) |  |
| RBC 3 |  | 1157 (64.7%) |
| Yes | 54 (4.6%) |  |
| No | 1103 (95.4%) |  |
| RBC 4 |  | 1165 (65.2%) |
| Yes | 51 (4.7%) |  |
| No | 1114 (95.7%) |  |
| RBC 5 |  | 1165 (65.2%) |
| Yes | 78 (6.6%) |  |
| No | 1087 (93.4%) |  |
| Amsterdam II Criteria |  | 1165 (65.2%) |
| Yes | 5 (0.4%) |  |
| No | 1160 (99.6%) |  |

First, missing data in the labels used for multitask supervision for *BRAF V600, KRAS* and Lynch Syndrome, were masked for loss computation. At each batch iteration, total loss was computed as a sum of losses for each supervision task and each of the task-specific losses was weighted by its corresponding ratio of missing data in the batch. This approach prevented batch fluctuations of total loss originated from variable amounts of missing points per batch.

Second, missing data in clinical variables used as inputs, were substituted with reserved values. Missing data in image inputs (including both tissue holes inside tiles as well as no image available) were imputed with the mean values of normalized images for each color channel. Segmentation of sub-tile tissue holes was done with thresholding operations based on the range of pixel values in the HSV colorspace.

Label imbalance, either in the primary task to be learned as well as in the number of tiles for each patient (which ranged from a minimum of 16 tiles to more than 2,000 tiles), was addressed with a composite weighted random sampling for both criteria. This method resulted in a training set of tiles that was class-balanced simultaneously for both the primary task label and the patient label. The primary task in experiments in Section 3.1 was the prediction of the MSI-status. *KRAS mut, BRAF V600 mut* and Lynch Syndrome were assigned primary task in some experiments described in Section 3.2 while in others were assigned auxiliary or secondary tasks.

### 2.5 Partitions, Data Augmentation, Model Training and Metrics

For model training, the dataset was split 80/20 for training and validation applying 5-fold cross-validation and guaranteeing on each fold that the images of each patient only belonged to one set (either training or validation but not both).

Tiles were resized to 224 *×*224 pixels and color was normalized following Mazenko method [30]. In addition to Mazenko, experiments were done with an additional color normalization with statistics computed from the EPICOLON image dataset which included 25 different hospitals obtaining equivalent results. Data augmentation at training and test time consisted in random rotations up to 90º, dihedral flips with probability of 0.5, a perspective warping of maximum 0.2, and hue variations of maximum 0.15. Of note, training sets included all magnifications so that the network could be trained simultaneously on higher tissue architecture patterns as well as on cellular-level features including nuclear characteristics. Models in experiments E3 to E6 (see Section 3.1) were trained for 3 epochs with batch size 512 while models leveraging a multiple instance learner based on transformers (E7-E9) were trained for one epoch only.

The statistical dependency between learned features and each of the selected biases was monitored during model training with the squared distance correlation *dc*. Principal component analysis (PCA) was used to assess how the spatial representations of the learned features were affected by the protected (bias) variables before and after bias ablation. Metrics used to assess the performance of the MSI classifier include ROC AUC as well as metrics determined from a chosen ROC threshold including balanced accuracy, sensitivity, specificity, positive predictive value, negative predictive value, false positive rate and false negative rates. The best threshold for each ROC curve was calculated as the point that maximized the difference between true positive rate and false positive rate. Metrics dependent on MSI-H prevalence are calculated assuming 15% in the real population.

### 2.6 Explainable Methods

SHAP (SHapley Additive exPlanations) [31] values were used to provide a means of visually interpreting the topology and morphology of features that influence predictions. The goal of SHAP is to explain the prediction of an instance by computing the contribution of each feature to the prediction. The SHAP explanation method computes Shapley values from coalitional game theory.

All code was programmed using the Python programming language. Machine Learning was implemented with Catboost library [29] and deep learning methods were implemented using FastAI [32] and PyTorch. QPath v0.2.3 [33] was used for annotation purposes.

## 3 Results

After the automatic filtering done by the preprocessing module M1, the final study sample totaled 1788 patients (171 MSI-High). The image dataset consisted of 1,065,479 tiles or adjacent regions without overlap including all magnifications and all types of tissues. The tissue classifier module M2 was then used in inference for the pre-selection of the regions of interest restricted to tumor epithelium, lymphocytic infiltrates and mucin totaling 523,624 tiles with 5 different magnifications.

This section describes the results of the different models trained on the dataset corresponding to those images and associated clinical data.

### 3.1 Experiments in MSI-status prediction

A total of 9 different models were trained in experiments (E1-E9) aimed to analyze the impact in MSI prediction accuracy from different input types, model architectures and various combinations of multitask supervision. The experiments and model results with detailed metrics are summarized in Figure 5 and Table 5.

**Table 4:** Revised Bethesda Criteria: Before the recommendation of universal screening testing, the revised Bethesda guidelines [2] were used to select patients for MSI testing. CRC tumors from individuals were recommended to be tested for MSI if any of the 5 criterion (RBC) were met. HNPCC: hereditary nonpolyposis colorectal cancer; MSI-H: microsatellite instability-high. * HNPCC-related tumors include colorectal, endometrial, stomach, ovarian, pancreas, ureter and renal pelvis, biliary tract, and brain (usually glioblastoma as seen in Turcot syndrome) tumors, sebaceous gland adenomas and keratocanthomas in Muir-Torre syndrome, and carcinoma of the small bowel. ** MSI-H like histology: Presence of tumor infiltrating lymphocytes. Crohn’s-like lymphocytic reaction, mucinous or signet-ring differentiation, or medullary growth pattern.

|  | **Revised Bethesda Criteria** |
| --- | --- |
| RBC 1 | Colorectal cancer diagnosed in a patient who is less than 50 years of age. |
| RBC 2 | Presence of synchronous, metachronous colorectal, or other HNPCC-associated tumors*, regardless of age. |
| RBC 3 | Colorectal cancer with the MSI-H like histology ** diagnosed in a patient who is less than 60 years of age |
| RBC 4 | Colorectal cancer diagnosed in a patient with one or more first-degree relatives with an HNPCC-related tumor, with one of the cancers being diagnosed under age 50 years. |
| RBC 5 | Colorectal cancer diagnosed in a patient with two or more first- or second-degree relatives with HNPCC-related tumors, regardless of age. |

**Table 5:**
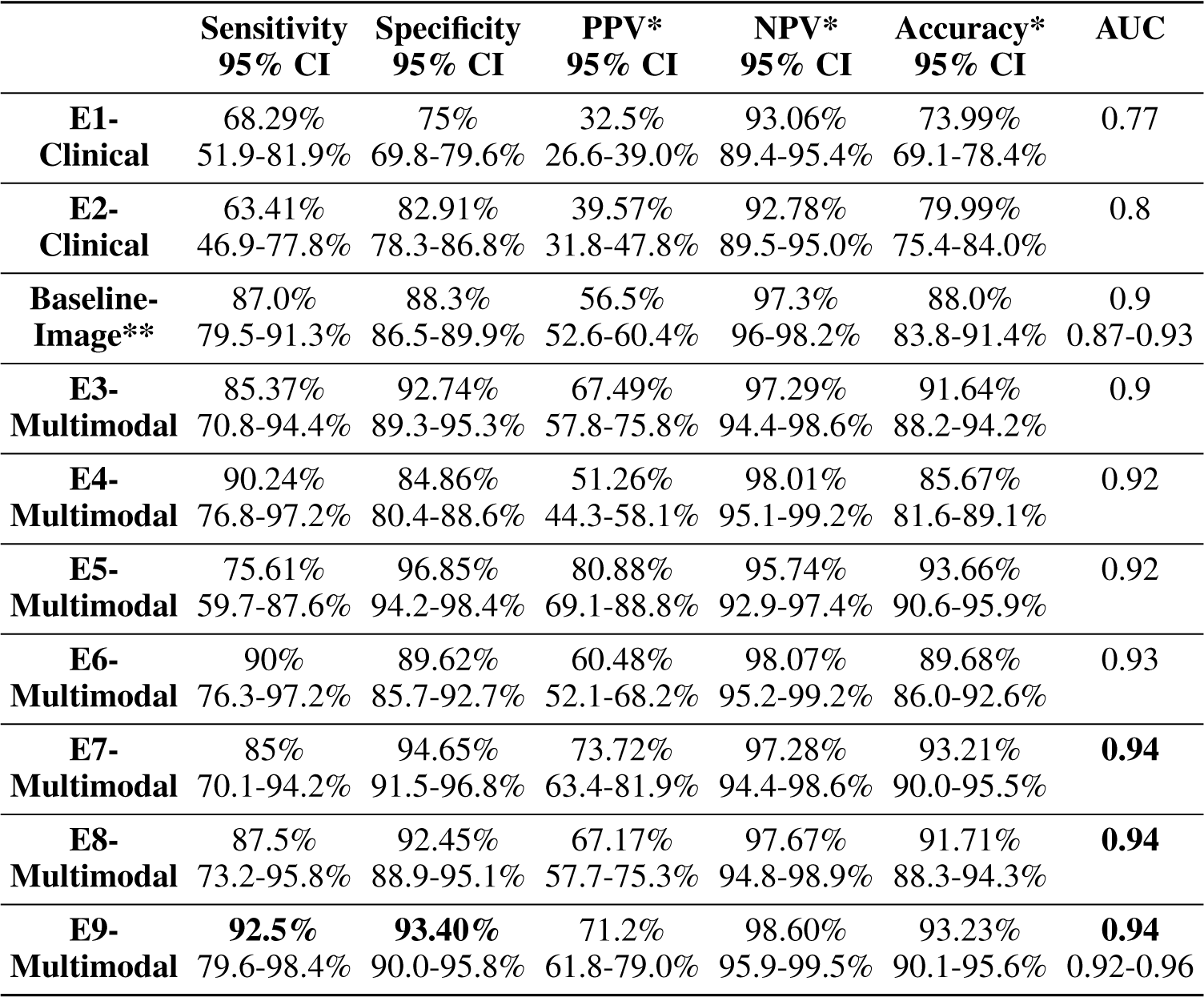
Patient level performance of the experiment models cross-validated in 5 folds for MSI status prediction. Metrics that are dependent on MSI-H prevalence, are calculated assuming a MSI-H prevalence in the real population of 15%. Confidence intervals for sensitivity, specificity and accuracy are “exact” Clopper-Pearson confidence intervals. Confidence intervals for the predictive values are the standard logit confidence intervals given by [34]. * Metrics that are dependent on MSI-H prevalence. ** xDEEP-MSI was used as baseline model trained on image only to compare with E3-E9 multimodal models trained with clinical and image inputs

|  | **Sensitivity**<br>**95% CI** | **Specificity**<br>**95% CI** | **PPV\***<br>**95% CI** | **NPV\***<br>**95% CI** | **Accuracy\***<br>**95% CI** | **AUC** |
| --- | --- | --- | --- | --- | --- | --- |
| **E1-Clinical** | 68.29%<br>51.9-81.9% | 75%<br>69.8-79.6% | 32.5%<br>26.6-39.0% | 93.06%<br>89.4-95.4% | 73.99%<br>69.1-78.4% | 0.77 |
| **E2-Clinical** | 63.41%<br>46.9-77.8% | 82.91%<br>78.3-86.8% | 39.57%<br>31.8-47.8% | 92.78%<br>89.5-95.0% | 79.99%<br>75.4-84.0% | 0.8 |
| **Baseline-Image\*\*** | 87.0%<br>79.5-91.3% | 88.3%<br>86.5-89.9% | 56.5%<br>52.6-60.4% | 97.3%<br>96-98.2% | 88.0%<br>83.8-91.4% | 0.9<br>0.87-0.93 |
| **E3-Multimodal** | 85.37%<br>70.8-94.4% | 92.74%<br>89.3-95.3% | 67.49%<br>57.8-75.8% | 97.29%<br>94.4-98.6% | 91.64%<br>88.2-94.2% | 0.9 |
| **E4-Multimodal** | 90.24%<br>76.8-97.2% | 84.86%<br>80.4-88.6% | 51.26%<br>44.3-58.1% | 98.01%<br>95.1-99.2% | 85.67%<br>81.6-89.1% | 0.92 |
| **E5-Multimodal** | 75.61%<br>59.7-87.6% | 96.85%<br>94.2-98.4% | 80.88%<br>69.1-88.8% | 95.74%<br>92.9-97.4% | 93.66%<br>90.6-95.9% | 0.92 |
| **E6-Multimodal** | 90%<br>76.3-97.2% | 89.62%<br>85.7-92.7% | 60.48%<br>52.1-68.2% | 98.07%<br>95.2-99.2% | 89.68%<br>86.0-92.6% | 0.93 |
| **E7-Multimodal** | 85%<br>70.1-94.2% | 94.65%<br>91.5-96.8% | 73.72%<br>63.4-81.9% | 97.28%<br>94.4-98.6% | 93.21%<br>90.0-95.5% | **0.94** |
| **E8-Multimodal** | 87.5%<br>73.2-95.8% | 92.45%<br>88.9-95.1% | 67.17%<br>57.7-75.3% | 97.67%<br>94.8-98.9% | 91.71%<br>88.3-94.3% | **0.94** |
| **E9-Multimodal** | **92.5%**<br>79.6-98.4% | **93.40%**<br>90.0-95.8% | 71.2%<br>61.8-79.0% | 98.60%<br>95.9-99.5% | 93.23%<br>90.1-95.6% | **0.94**<br>0.92-0.96 |

#### 3.1.1 E1-Clinical Model

A Catboost MSI-status classifier was trained (with hyperparameters as described in Section 2.3) using as inputs only three variables: gender, tumor localization and age at primary CRC diagnosis. The AUC reached 0.77 and sensibility and specificity with 95% CI was 68.29% (51.9%-81.9%), and 75% (69.8%-79.6%) respectively.

#### 3.1.2 E2-Clinical and Hereditary Model. Explainability and Priorization of Variables

A similar model was trained as E1-Clinical but adding oncology-family history variables as inputs (see Table 3). The AUC increased from 0.77 to 0.8, sensibility decreased to 63.41% (46.9%-77.8%) and specificity increased to 75% (69.8%-79.6%).

The seven most relevant features for classifying the samples as MSI-high vs MSS were, by decreasing order of importance, age at diagnosis (being more pronounced below 60 years), localization of CRC (right-sided), gender (female), synchronous adenoma, number of family member with HPNCC-related tumor, number of family member with CRC and meeting the revised Bethesda Criteria (see Fig. 4).

**Figure 4:**
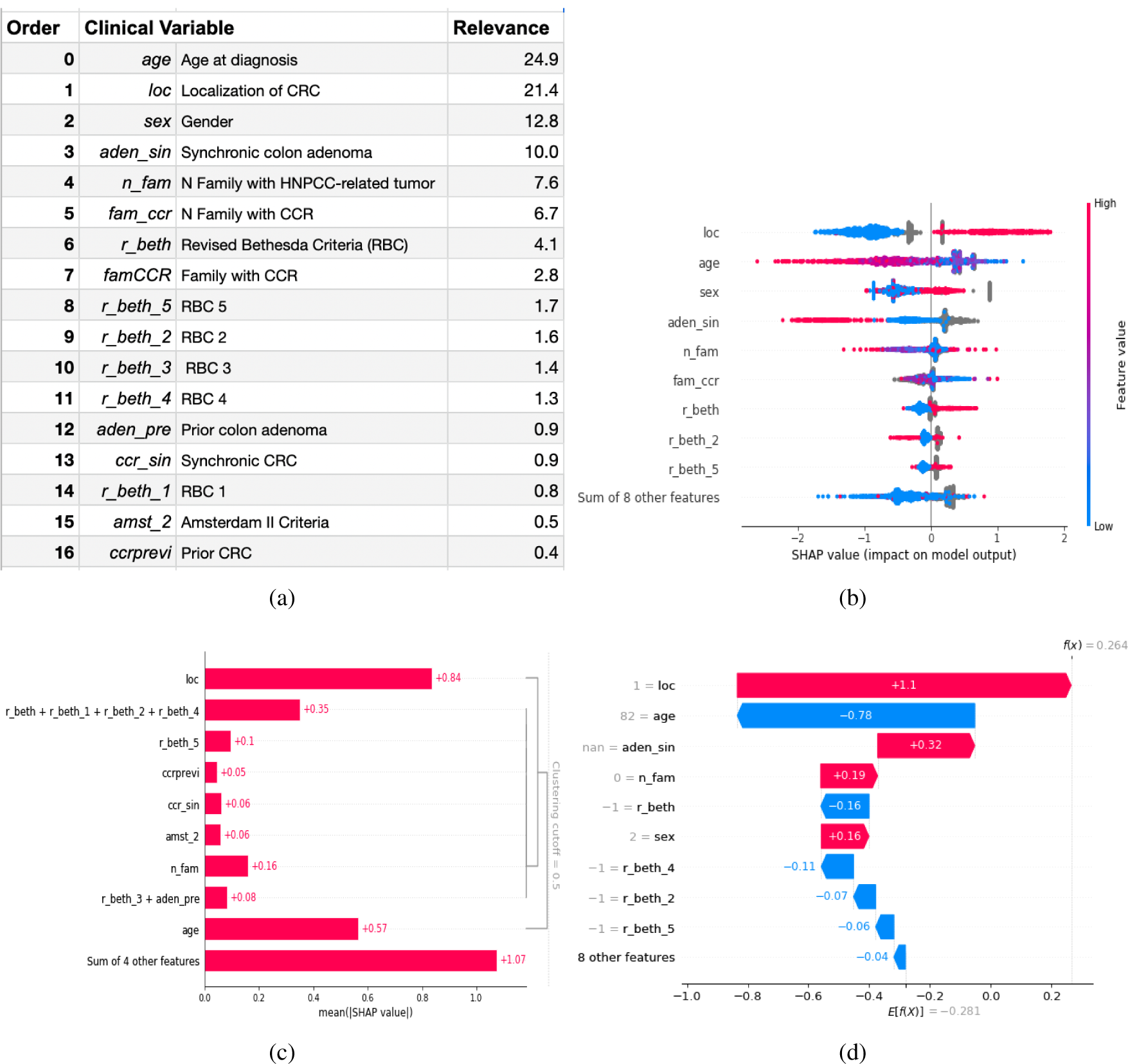
Visual explainability of clinical variables. Figure (a) shows all clinical variable ordered by their impact on the prediction of MSI-status. Figure (b) shows the distribution of the impacts each variable has on the MSI-status. Variables are sorted from top to bottom by the sum of SHAP value magnitudes over all samples. The color represents the variable value: for continous variables red denotes high values and blue low, for binary variables red denotes Yes and blue No, for localization of CRC red is right-sided and blue otherwise and for sex red denotes females and blue males. Figure (c) shows variables hierarchically clustered based on redundancy. Variables are ordered by relevance in each cluster. Figure (d) explains how each of the variables contributed to the output for a given patient. Variables pushing the MSI status prediction towards MSI-H are shown in red while those influencing it towards MSS are in blue. On this patient’s case, being a woman diagnosed with right-sided CRC shifted the prediction towards MSI-H, while being elderly (82 years) and not meeting any of the revised Bethesda criteria shifted the outocome towards MSS, yielding a net positive decision towards MSI-H.

We measured feature redundancy through model loss comparisons, building a hierarchical clustering of the features by training separate models to predict the outcome for each pair of input features. Figure 4c shows the variables organized in hierarchical clusters with distance < 0.5 being 0 perfectly redundant variables and 1 independents. As expected, the cluster with highest redundancy included the revised Bethesda Criteria (RBC) and its related variables where the number of family members with HNPCC-related tumors was the single variable with the most impact in the group. Right-sided localization was the next closest variable to the RBC cluster followed by age, and their high impact on predictions makes them the two more informative and less redundant variables.

#### 3.1.3 E3-Multimodal Model

Next, after analyzing in prior section the relative impact and redundancy between clinical variables, a multimodal DL-model was trained on the three most informative clinical variables (age, tumor localization and gender) and image of CRC tissue. The model achieved a AUC of 0.9 (at patient-level by majority vote), and 85.37%(70.8-94.4%) 92.74%(89.3-95.3%) of sensitivity and specificity respectively. As expected this model increased all metrics over models trained in clinical information only (E1 - E2). On the other hand, when compared with the baseline model *xDEEP-MSI* which was trained on image only, the specificity increased from 88.3% to 92.7% while reaching the same AUC in 0.9.

#### 3.1.4 E4-Multimodal Multitask Model

On this experiment the same model described in E3 was trained adding multitask supervision including MSI-status, *KRAS* and *BRAF V600*. Missing values for *KRAS* and *BRAF V600* were masked for loss computation and task-specific losses were weighted by missing frequency as described in Section 2.4. This approach increased both the AUC (at patient-level by majority vote) from 0.9 to 0.92 and the sensitivity from 85.37% to 90.24% as a result of a lower number of false negatives. Nonetheless the model decreased specificity to 84.86% from a relative increase in false positives.

#### 3.1.5 E5-Multimodal Multitask Model

Next, all family and oncology variables as described in Table 1 were inputed together with age, localization, gender and image. In the same manner as described in E4, the model was trained with multitask supervision in MSI-status, *KRAS* and *BRAF V600*. This approach, similarly as observed in E2, where family and oncology variables were added, contributed to increase specificity up to 96.85% while decreased sensibility and the AUC 0.92 (at patient-level by majority vote) remained unchanged as compared with E4.

#### 3.1.6 E6-Multimodal Multitask Model with Patient-attention

This experiment built upon E5 by adding Module 4 (see Section: 2.2.4), a multiple instance learner with attention-mechanisms based on transformers with a per-patient output. The system was similarly trained on the same multimodal inputs (age, tumor localization, gender, oncology-family history variables and CRC tissue images) and was supervised on MSI-status, *KRAS* and *BRAF V600*. This approach further improved the AUC to 0.93 and yielded both high sensitivity and specificity.

#### 3.1.7 E7-Multimodal Multitask Model with Patient-attention and Stochastic Sampling

This experiment was similar as E6 with two differences. First, sampling of tiles by patient inputted to the transformer applied an stochastic random sampling of *n* = 8 tiles. Second, the magnification level for each tile was inputted in the model (see Section 2.2. The AUC increased to 0.94

#### 3.1.8 E8-Multimodal Multitask Model with Patient-attention and Stochastic Sampling

This experiment built upon E7 model but added supervision for the composite variable *KRAS mut* or *BRAF V600 mut*, in addition to MSI-status, *KRAS mut* and *BRAF V600 mut* separately. The AUC remained 0.94.

#### 3.1.9 E9-Multimodal Multitask Model with Patient-attention and Stochastic Sampling

This experiment built upon E8 model but added supervision for confirmed Lynch syndrome, in addition to MSI-status, *KRAS* and *BRAF V600* separately and the composite variable *KRAS mut* or *BRAF V600 mut*. While the AUC remained 0.94, both sensitivity and specificity in the best ROC threshold, were above 90%, with 92.5% and 93.4% respectively.

Summarizing, as shown in Fig. 5 and Table 5, the performance in the prediction of MSI-status steadily increased from using only age at diagnosis, localization of CCR and gender (AUC 0.77), adding oncology and family variables (AUC 0.8), adding H&E tumor slide images (AUC 0.9), adding multitask supervision for the presence of *KRAS* and *BRAF V600* (AUC 0.92), adding a self-supervises multiple instance learner with patient-attention (AUC 0.93), adding stochastic sampling of tiles and additional multitask supervision for confirmed Lynch syndrome (AUC 0.94)

**Figure 5:**
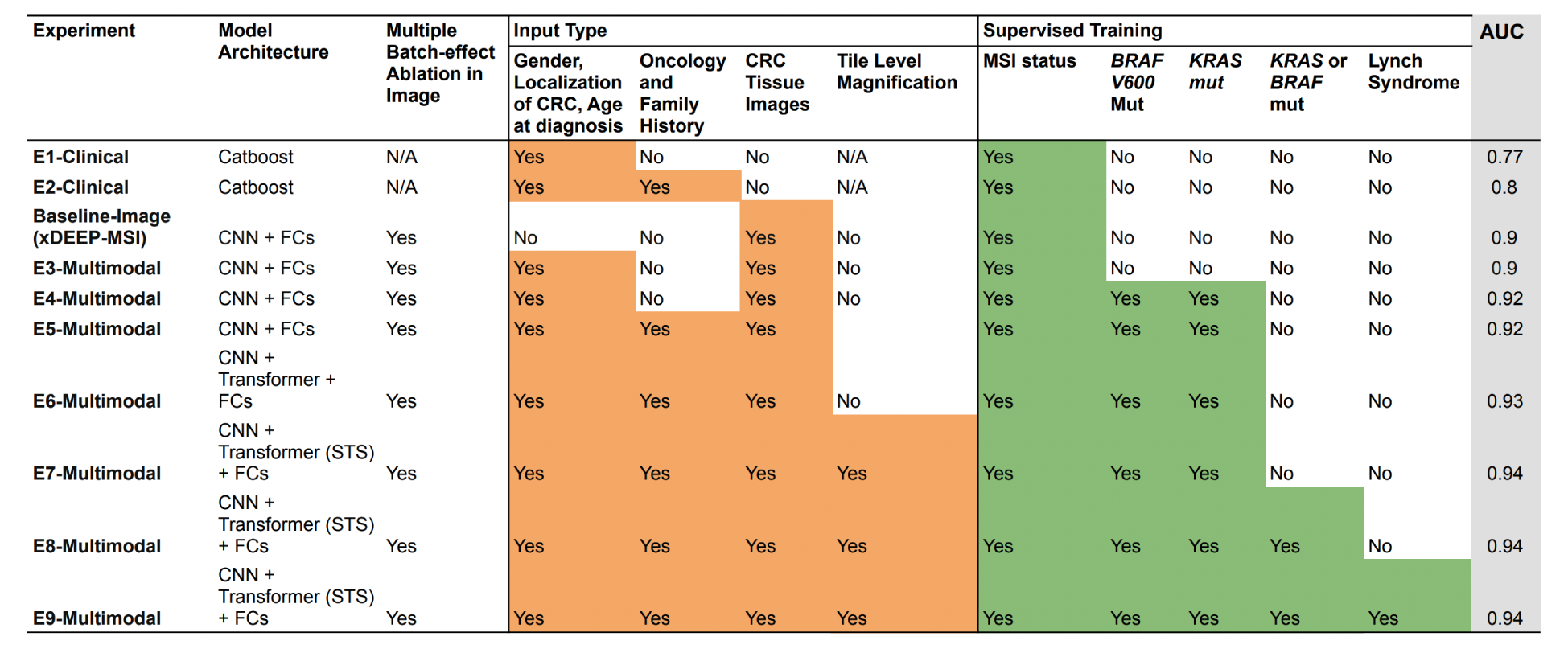
Model Training Experiments: A summary of the differences between models from the different experiments regarding architecture, input types and molecular supervision tasks used for training. Model architecture lists all elements trained end-to-end. *STS*: Stochastic tile sampling for selecting the patient’s set of *n* tiles imputed in the transformer. *CNN*: Convolutional Neural Network used as backbone in M3. *FCs*: Fully connected neural layers used in M5 and M6. *Transformer*: Encoder-Decoder Neural Network based on attention mechanisms used in M4.

### 3.2 *KRAS mut, BRAF V600 mut* and Lynch Syndrome predictions

The best models on the primary task of MSI-status (E7-E9) were tested for performance in *KRAS*, textitBRAF, composite variable *KRAS mut* or *BRAF V600 mut* and Lynch Syndrome prediction. All of those tasks were considered secondary auxiliary tasks given 1) the extreme imbalance and scarce number of Lynch Syndrome cases (with 0.8% prevalence in the final study sample) and 2) the high missing rate (above 75%) for *KRAS mut,BRAF V600 mut*, composite variable *KRAS mut* or *BRAF V600 mut*. The AUC of the E9 Model on the prediction of Lynch Syndrome validated on a sample with only 3 cases from a total of 322 patients was 0.62. The AUC of E8 model on the prediction of *BRAF V600 mut, KRAS mut* and composite variable *KRAS mut* or *BRAF V600 mut* was respectively 0.73, 0.57 and 0.71. Moreover in order to try improving those results we trained independent models similar to E9 (see Section 3.1.9) but handling each of those variables as primary task where the sampling was weighted by either *BRAF V600 mut* or *KRAS mut* frequencies instead of *MSI-H* frequency. The AUC for the prediction of *BRAF V600 mut* and *KRAS mut* increased to 0.77 and 0.62 respectively.

As summary, all of these predictive molecular tasks were consistently under-performing in all the different experiments done, with metrics below 0.8.

### 3.3 Effect of incomplete data at inference time for MSI status prediction

Next, we analyzed the effects in model’s MSI-status predictive performance by varying combinations of missing multimodal inputs at inference. Model E8 3.1.8 which was regarded as one of the best multimodal model, was chosen for testing the different combinations of inputs on inference.

The DL system, specifically Module 5 (see 2.2.5), allows different combinations of missing data to be entered depending on their availability for each patient case. Imputation of missing data for the different data modalities (digitized image of tumor tissue, general clinical variables and oncological-family history variables) was done as explaind in Section 2.4.

The highest performance with an AUC of 0.94 was obtained by entering all types of multimodal data (image, general clinical variables, oncology and family history), followed by an AUC of 0.93 entering only image and general clinical variables (age at diagnosis, CRC localization and gender), followed by an AUC of 0.9 entering only image, followed by an AUC of 0.76 only using general clinical, oncology and family history data, followed by an AUC of 0.74 only using age at diagnosis, tumor localization and gender. Also as it can be observed in Fig 5 the trend and level of performance in inference for the best performing model matched the AUC results from E3-E9 models, each trained on the same respective combinations of input modalities. This indicates that a single model could be used for varying completeness of data in different clinical scenarios dictated by the availability of variables.

As conclusion, the degree of completeness in input data impacted the performance of predictions of the model in inference and whenever the image was included the results in inference were equal or above 0.9 of AUC, reaching up to 0.94 when all clinical variables were entered.

Lastly, an important system parameter for inference is the number *n* of tiles chosen with stochastic random sampling (STS) which are inputted in the patient-attention module M4 (see Section 2.2.4) as a sequence of length *n* where each element is a feature representation of each tile. In order to decide the optimal number *n* Module 7 (see 2.2.7) was invoked in inference mode applying test-time image augmentations, with *m* = 1 while increasing values of *n*. We observed a steady gain in performance from *n* = 8, 16, 32, and 40, without further improvements for larger values while maintaining constant *m* = 1. For all inference runs detailed above, we used *m* = 1 and *n* = 40, which means that for a given patient a prediction was obtained from a total of 40 tiles. On the other hand parameter *m* when assigned values above 1 produce an statistical distribution of predictions by patient where each prediction is based on the stochastic sampling of *n* tiles.

## 4. Discussion

We present a new system for the prediction of MSI from H&E images using artificial vision techniques that builds upon that published in [18] with the goal to increase its capabilities by leveraging clinical multimodal-inputs, multitask-molecular weekly supervision and self-supervised multiple instance learning at patient level. The new functional modules are adapted to the prior system which consisted in an end-to-end TMA-customized image preprocessing module to tile samples at multiple magnifications in the regions of interests guided by the automatically detected type of tissues and a multiple bias distiller system integrated with the MSI predictor.

The highlights of this work are as follows:

First, the present work leverages multimodal data consisting of a rich selection of informative clinico-pathology variables for MSI-status ascertainment: pathology images of tumor tissue (including its magnification level), clinical variables (age, localization of colorectal cancer and gender), hereditary information and oncological history (revised Bethesda criteria, Amsterdam II criteria, number of family members with colorectal cancer, number of family members with any HPNN tumors, synchronic colorectal cancer, previous colorectal cancer, synchronic colorectal adenomatous polyps and previous colorectal adenomatous polyps. We analyzed the contribution of each the data types in the model’s performance for MSI-status prediction. The highest performance with an AUC of 0.94 was obtained by entering all types of multimodal data (image, general clinical variables, oncology and family history), followed by an AUC of 0.93 entering only image and general clinical variables (age at diagnosis, CRC localization and gender), followed by an AUC of 0.9 entering only image, followed by an AUC of 0.76 only using general clinical, oncology and family history data, followed by an AUC of 0.74 only using age at diagnosis, tumor localization and gender. As expected, when only tabular data was entered (without image inputs), the benchmark based on a Catboost model 3.1.2 had superior performance reaching an AUC of 0.8 using general clinical, oncology and family history data. Importantly, the most informative and less redundant clinical variables were age, CRC localization and gender, which is also the most easy and readily available information in the clinical practice. Additionally, in agreement with the literature, young ages at diagnosis, right localization of the CRC and female gender were the main factors increasing the odds of MSI-H status. Our findings suggest that whenever digital tumor images are available a single DL-model could be used for varying completeness of data in different clinical scenarios dictated by the availability of variables. The degree of completeness in input data impacted the performance of predictions of the model in inference and whenever the image was included the results in inference were equal or above 0.9 of AUC, reaching up to 0.94 when all clinical variables were entered. In clinical practice, clinico-pathological information is not always available for a given patient, hence the utility of a system could be maximized if it is designed to handle missing or incomplete information.

Second, the new system is designed to predict different molecular alterations simultaneously, MSI status, *KRAS mut,BRAF V600 mut*, composite variable *KRAS mut* or *BRAF V600 mut* and Lynch Syndrome. Nonetheless only MSI (ROCAUC 0.94) reaches clinical-level performance (i.e. specificity and sensitivity is above 0.8). All other tasks, *KRAS mut,BRAF V600 mut*, composite variable *KRAS mut* or *BRAF V600 mut* and Lynch Syndrome prediction are considered in the present work as secondary auxiliary tasks that proved to increase MSI-status prediction performance as shown in Table 5 experiments E3 to E9. It is widely accepted that multitask learning, where a single model learns multiple tasks simultaneously, is able to learn a more general representation helping to avoid model over-fitting, providing implicitly a technique of data augmentation and helping to focus on relevant features in the presence of noisy labels. In line with it, we demonstrated that multitask supervision yielded a more robust model with increased performance in the primary task of MSI status prediction.

Third, in an effort to make the model learn the most relevant tissue areas with the strongest signal-to-noise ratio for the different molecular alterations we leverage a self-supervised multiple instance learning module (2.2.4 based on a encoder-decoder transformer architecture to integrate the tile information at patient level. It also introduces a per-patient stochastic sampling of tiles. Specifically, the DL-system applies an iterative stochastic sampling of tiles by patient, learns the image features of each tile and integrates them in a transformer architecture at patient level, extracting a patient level latent representation that is then fed to the multitask and multiple bias classification heads. Training the model at patient level instead of at tile level, allows it to learn which tiles have the strongest signal for each molecular alteration. We propose this architecture, similar to [35] who also applied attention-based multiple instance learning, to overcome the limitations of noisy labels introduced by weak supervision and it is an alternative to the one proposed in [19] that used an iterative draw and rank sampling method to help prioritize the tiles with the strongest signal. We show that our approach, even in a different dataset, obtains results in line with those reported by [19] for each of the different molecular alterations tested: mean AUROCs for the prediction of microsatellite instability (0.94 vs 0·86 [0·04]), BRAFmut (0·77 [0·01] vs 0·79 [0·01]), and KRASmut (0·62 [SD 0·04] vs 0·60 [SD 0·04]).

Summarizing, the performance in the prediction of MSI-status steadily increased from training models using only age at diagnosis, localization of CCR and gender (AUC 0.77), adding oncology and family variables (AUC 0.8), adding H&E tumor slide images (AUC 0.9), adding multitask supervision for the presence of *KRAS mut* and *BRAF V600 mut* (AUC 0.92), adding a patient-attention layer with transformers (AUC 0.93), adding stochastic sampling of tiles by patient in the patient-attention layer and additional multitask supervision for confirmed Lynch syndrome (AUC 0.94)

The present system applies also the same methodology as in xDEEP-MSI. Specifically 1) it addresses the multiple batch effects inherent in the dataset (project of origin, patient and TMA-glass bias) with an adversarial multiple bias ablation technique. Similarly we verify the adequate ablation of batch-effects by monitoring the distance correlation between the extracted features of the images and each of the biases during training. 2) Not only tumor epithelium but also the mucine and lymphocytic infiltrate regions were included, as those regions were probed in [18] to be nonspecific at image level, but to increase the sensibility at patient level, which is a desirable characteristic for screening purposes. 3) It includes 5 different magnifications (*×* 0, *×* 5, *×* 10, *×*20, *×* 40), helping the model to learn both low and high level tissue architectural patterns at the same time.

Limitations of the study are as follow: In the final study sample after image pre-processing the resulting frequencies for the molecular characteristics, in particular for MSI-H, as discussed in [18] and Lynch Syndrome cases, were lower than those of the original EPICOLON population and/or those expected in the clinical practice. To approximate models performance in the clinical setting, metrics which are impacted by disease prevalence, are calculated considering the MSI-H prevalence in the real population (15%). Considering together the EPICOLON and HGUA projects (see Table 2), there was a high missing rate (above 75%) for *KRAS mut,BRAF V600 mut*, composite variable *KRAS mut* or *BRAF V600 mut*, and extreme label imbalance with scarce number of Lynch Syndrome cases available (0.8% prevalence in the final study sample). In fact, there where only 13 cases of Lynch Syndrome and 40 positive cases of *BRAF V600 mut* available. The AUC for the prediction of Lynch Syndrome and *BRAF V600 mut* was 0.62 and 0.77 respectively. The AUC for the prediction of *KRAS mut* was 0.62. All those predictive molecular tasks were consistently unsatisfactory in all the different experiments done, with metrics below 0.8. We hypothesize that predicting Lynch Syndrome and *BRAF V600 mut* from H&E tissue slide images are potentially feasible tasks for deep learning methods with a larger number of positive cases. Conversely, *KRAS mut* prediction is a much more challenging task and even with larger sample sizes (115 positive cases on this study) the low performance achieved, in line with other works [19]) below 0.65, may be interpreted as predictions by chance or just learning of non-informative statistics such as the label frequency. This finding suggest that the signal is too weak to be learned from the H&E stained images and additional research is still needed.

As future work, testing the generalizability of the system in an independent and prospective test would be necessary.

## 5 Conclusions

In this study, we present a new AI system for the prediction of MSI in colorectal cancer from H&E images in TMAs that builds upon xDEEP-MSI enriching it with 1) multitask supervised learning for multiple molecular alterations (MSI status, K-RAS and BRAF mutations and Lynch Syndrome confirmed by germline mutations), 2) multimodal inputs tolerant to variable degree of completeness that includes image, age, gender, localization of CRC, revised Bethesda criteria, Amsterdam II criteria and additional oncological history and 3) self-supervised multiple instance learning with per patient attention-mechanisms to obtain patient-level predictions. The AUC at patient level, and including all three selected tissues (tumor epithelium, mucin and lymphocytic regions) and 5 magnifications, increases from 0.9 ± 0.03, as previously reported, to 0.94 ± 0.02.

To the best of our knowledge this is the first work that uses multimodal inputs and multiple molecular supervision for the prediction of MSI in CRC from H&E. Prospective validation in an external independent dataset is still required.

## Supporting information

Author's COI Disclosure

Author's COI Disclosure

Author's COI Disclosure

Author's COI Disclosure

TRIPOD-CHECKLIST

## Data Availability

The data from EPICOLON project and HGUA
are not publicly available, in accordance with the research group and institutional requirements governing human subject privacy protection. This project was approved by the institutional research committee CEIm PI2019-029 from ISABIAL and both the
images and associated clinical information was previously anonymized.

## Abbreviations

The following abbreviations are used in this manuscript:

BE: batch effect module learners
FE: feature extractor
MSI: Microsatellite Instability
dc: squared distance correlation

## CRediT authorship contribution statement

**Aurelia Bustos**: Conceptualization, Methodology, Software, Validation, Formal analysis, Resources, Writing - Original Draft, Visualization. **Artemio Payá**: Conceptualization, Writing - Review & Editing, Data Curation.**Andres Torrubia**: Methodology, Software, Resources, Writing - Review & Editing. **Cristina Alenda**: Conceptualization, Investigation, Resources, Data Curation, Writing - Original Draft, Writing - Review & Editing, Supervision, Funding acquisition

## Acknowledgements

We want to particularly thank the patients and the BioBank ISABIAL integrated in the Spanish National Biobanks Network and in the Valencian Biobanking Network for their collaboration.

## CRediT authorship contribution statement

Conceptualization, C.A. and A.B. and A.P.; methodology, A.B. and C.A. and A.T.; software, A.B. and A.T.; validation, A.B. and C.A.; formal analysis, A.B.; investigation, A.B. and A.P. and A.T. and C.A.; resources, C.A. and A.B.; data curation, C.A.; writing—original draft preparation, A.B. and C.A.; writing—review and editing, A.B. and A.P. and A.T. and C.A.; visualization, A.B.; supervision, C.A.; project administration, C.A.; funding acquisition, C.A. All authors have read and agreed to the published version of the manuscript.

## Institutional Review

The study was conducted according to the guidelines of the Declaration of Helsinki, and approved by the Institutional Review Board of ISABIAL (CEIm PI2019-029).

## Informed Consent

Informed consent was obtained from all subjects involved in the study.

